# Trajectories of Ankle-Brachial Index Values and Their Relation to Cardiovascular Health Measured by Life’s Essential 8 in the Atherosclerosis Risk in Communities Study

**DOI:** 10.64898/2026.07.20.26358527

**Authors:** Sydney E. Browder, Corey A. Kalbaugh, Katharine L. McGinigle, Kelly R. Evenson, Sara Jones Berkeley, Wayne Rosamond

## Abstract

**Background:** Peripheral artery disease (PAD) is an occlusive arterial disease primarily affecting the lower extremities. It impacts over 230 million people worldwide and is associated with significant morbidity and mortality. The ankle brachial index (ABI) test is a non-invasive method to detect PAD that compares the blood pressure in the ankle and arm to evaluate lower extremity blood flow. An estimated 20-50% of individuals with detectable PAD are asymptomatic and remain undiagnosed; however, ABI screening in high-risk, asymptomatic populations is not currently guideline-recommended. Few studies have evaluated change in ABI over time in asymptomatic populations. Therefore, we aimed to identify distinct trajectories of ABI values from mid-to late-life.

**Methods:** We utilized data from the Atherosclerosis Risk in Communities (ARIC) study; a longitudinal cohort study initiated in 1987 that enrolled 15,792 participants aged 45-64. ABI measurements were collected at five visits over a 30-year period. We used group-based trajectory modeling to identify trajectories of ABI from mid-to late-life. Final model selection was based on visual fit, statistical criteria, group sizes, and substantive knowledge. Lastly, we compared baseline demographics, social determinants of health, and overall cardiovascular (CV) health, assessed using the American Heart Association’s Life’s Essential 8 (LE8) framework, across trajectory groups.

**Results:** We identified 4,121 participants with ≥3 ABI measurements over the study period in at least one limb. At baseline, participants had an average age of 51.4 ± 4.9 years, were 57.3% female, 22.2% Black, and had an average overall LE8 score of 68.0 ± 13.9 points. Our final model identified three linear trajectories: low-normal, high-normal, and declining. Overall LE8 scores varied significantly across trajectory groups: 67.3 ± 10.7 (high-normal), 61.9 ± 13.3 (low-normal), and 50.1 ± 15.8 points (declining). Women had lower average ABI values, were more likely to experience a declining ABI trajectory, and had a delayed onset of decline compared to men. A greater proportion of Black participants experienced declining ABIs, with earlier, faster, and more severe declines than White participants.

**Conclusions:** Poor overall CV health and common CV risk factors are associated with ABI decline. Targeted ABI screening in middle age may help detect PAD in its beginning stages and support early intervention.

## Introduction

Peripheral artery disease (PAD) affects more than 230 million people worldwide and is the leading cause of lower extremity amputations in the US.^1,2^ PAD is characterized by stenosis of the arteries in the extremities, primarily the legs, caused by the build-up of atherosclerotic plaque.^3^ Symptoms include claudication, rest pain, ischemic wounds, and gangrene, which can quickly progress to amputation or death.^4^ Although traditional PAD treatments have improved in recent decades, the global prevalence, incidence, and deaths related to PAD increased by 73%, 72%, and 146%, respectively, from 1990 to 2019 and the global number of PAD cases are projected to increase by 220% by 2050.^5,6^ These trends underscore that, while improvements in treatment are essential, efforts to reduce the population burden of PAD will require *earlier identification of PAD* and *better risk factor control*.

When identified early, PAD can often be managed with lifestyle modifications, pharmacotherapy, and structured exercise programs; however, advanced disease typically requires invasive interventions (e.g., revascularization or amputation). The ankle brachial index (ABI) test is a non-invasive, inexpensive test used to detect PAD by comparing the blood pressure in the ankle to that in the arm.^7^ The American Heart Association recommends patients presenting with a suggestive clinical history or abnormal physical examination of the lower extremities receive prompt evaluation with an ABI test;^8^ however, it has been estimated that 20-50% of individuals with detectable PAD are asymptomatic and, therefore, often remain undiagnosed until they develop symptoms.^9^ Although the severity of chronic limb-threatening ischemia, the worst form of PAD, has been likened to terminal cancer, there are currently no recommended screening guidelines for PAD using the ABI test in high-risk, asymptomatic individuals.^10^

In 2018, the US Preventive Services Task Force (USPSTF) concluded the available evidence was *insufficient* to assess the benefits and harms of ABI screening in asymptomatic adults^11^ The natural progression of ABI values from mid-to late-life, when PAD typically develops, remains poorly understood, as only five studies have evaluated change in ABI over time in asymptomatic populations.^12–16^ These studies varied in sample size, age, follow-up, and ABI measurement frequency and none conducted a formal trajectory analysis of ABI progression from mid-life into late-life. To better understand the natural progression of ABI values, this study aimed to identify distinct trajectories of ABI values from mid-to-late life and quantify the baseline cardiovascular (CV) health within each trajectory group using the American Heart Association’s Life’s Essential 8 (LE8) framework among participants from the Atherosclerosis Risk in Communities (ARIC) study.

## Methods

### Study Design and Population

The ARIC study is a longitudinal, prospective cohort study investigating the etiology of atherosclerosis and CV disease that began in 1987.^17^ ARIC investigators randomly selected 15,792 participants from four ARIC communities (suburban Minneapolis, MN; Forsyth County, NC; Washington County, MD; and Jackson, MS) to participate in the ARIC cohort. At baseline, participants completed a comprehensive evaluation that included demographic, clinical, and social assessments, as well as CV measurements and standardized questionnaires. We chose the ARIC cohort as the study population for this project due to the availability of repeat ABI measurements over multiple decades and collection of variables necessary to calculate LE8 scores at baseline. Our 30-year study period ranges from ARIC Visit 1 (1987-1989) to Visit 7 (2018-2019).

### Study Sample

Non-White and non-Black participants (n = 48 participants), Black participants from Minneapolis and Washington Co. (n = 55), limbs with <3 ABI measurements (n = 26,182 limbs), and limbs receiving a lower extremity revascularization procedure before three ABI measurements (n = 18 limbs) were excluded from our study sample, resulting in 4,121 participants with 5,178 qualifying limbs. For participants with two qualifying limbs, one was randomly selected as the index limb to ensure independent observations. This index limb was the followed throughout the study period. The final analytic sample consisted of 4,121 participants, each contributing one index limb.

### ABI Measurements

The ABI test utilizes the ratio of systolic blood pressure in the ankle compared to the systolic blood pressure in the brachial artery of the arm to assess blood flow through the lower extremities. ABIs are continuous values that are frequently categorized as arterial calcification (>1.4), normal (1.0-1.4), pre-PAD (0.9-<1.0), PAD (0.5-<0.9), and chronic limb-threatening ischemia (<0.5). In the ARIC study, resting ankle and brachial systolic blood pressures were collected on all available participants at Visits 1 (n = 15,220, 96.4%), 5 (n = 5,610, 85.8%), and 7 (n = 2,872, 80.0%) and on a simple random sample of the cohort at Visits 3 (n = 4,575; 35.5%) and 4 (n = 6,404; 54.9%). Ankle pressure was measured in <u>one</u> limb at Visits 1, 3, and 4 and in <u>both</u> limbs at Visits 5 and 7. Detailed descriptions of ARIC blood pressure collection methods and ABI calculations are available in ARIC manuals 6a and 11.^18^

### Covariates

Demographic covariates included baseline continuous age (years), sex (male/female), self-reported race (Black = B or White = W), and ARIC study site (Forsyth County = F, Jackson = J, Minneapolis = M, Washington County = W). Due to collinearity between race and study site, we combined race and ARIC study site into a five-level race-center categorical variable (BF, BJ, WF, WM, WW). We did not observe collinearity between any other covariates. Other covariates included access to care (poor/moderate/good), history of CV disease (yes/no), individual socioeconomic status (SES; low/average/high), mental health (poor/good), neighborhood SES (low/average/high), social support (poor/moderate/good), and continuous overall and individual [blood glucose, blood pressure, body mass index (BMI), cholesterol, diet, physical activity, sleep, and smoke] LE8 scores. Most of these covariates were collected at ARIC Visit 1 (baseline), with the exception of blood glucose, mental health, sleep, and social support which were collected at Visit 2 and used as a proxy for Visit 1. We calculated overall and component LE8 scores based on previously established criteria.^19,20^ Briefly, each of the eight LE8 components was scored 0-100 points and averaged to produce an overall LE8 score (0-100 points). Higher LE8 scores represent better CV health; 0-49 points is considered poor, 50-79 points is moderate, and 80-100 points represents ideal CV health. We identified lower extremity revascularizations across the study period using relevant ICD-9 and ICD-10 codes. These codes, plus additional information on how we operationalized covariates from ARIC data, are available in Supplemental Table 1.

### Selection Bias and Missing Data

The exclusion criteria reduced our sample from 15,792 to 4,121 participants. Excluded individuals were older at baseline, more often male and Black, had poorer social determinants of health, were more likely to have a history of CV disease and revascularization, and had worse overall and component LE8 scores (Table 1). Excluded limbs were also more likely to have ABI values in the PAD range (0.5-<0.9) across visits (Supplemental Table 2). These differences between included and excluded participants indicated the presence of selection bias. To address this, we be built stabilized inverse probability weights (IPWs) to better represent the overall ARIC population. We modeled the bivariate association between each covariate and inclusion in our study using logistic regression to determine which variables to include in our IPWs. Baseline age, history of CV disease, healthcare access, individual SES, neighborhood SES, mental health, overall LE8, race-center, sex, social support, and revascularization were associated with inclusion.

**Table 1.**
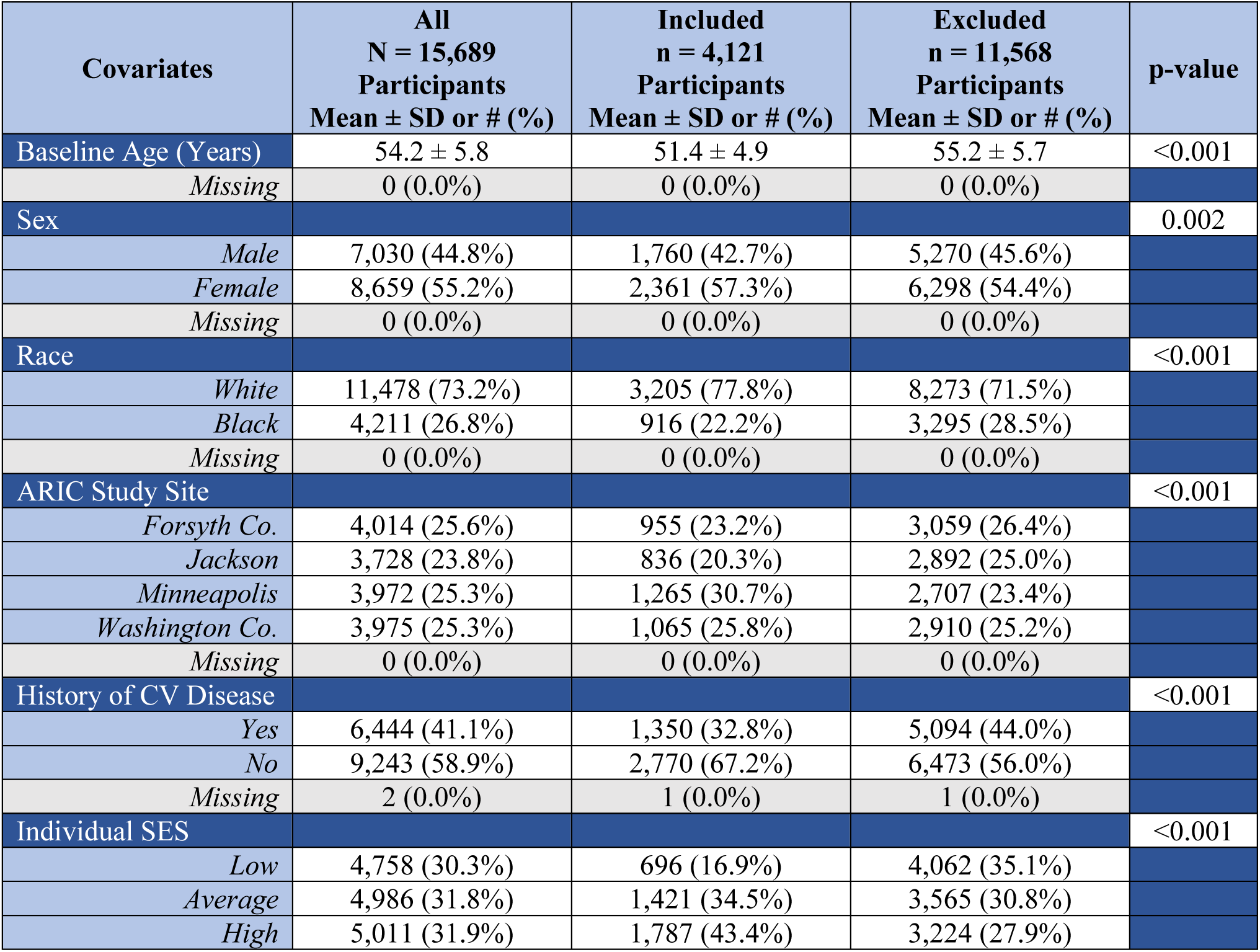
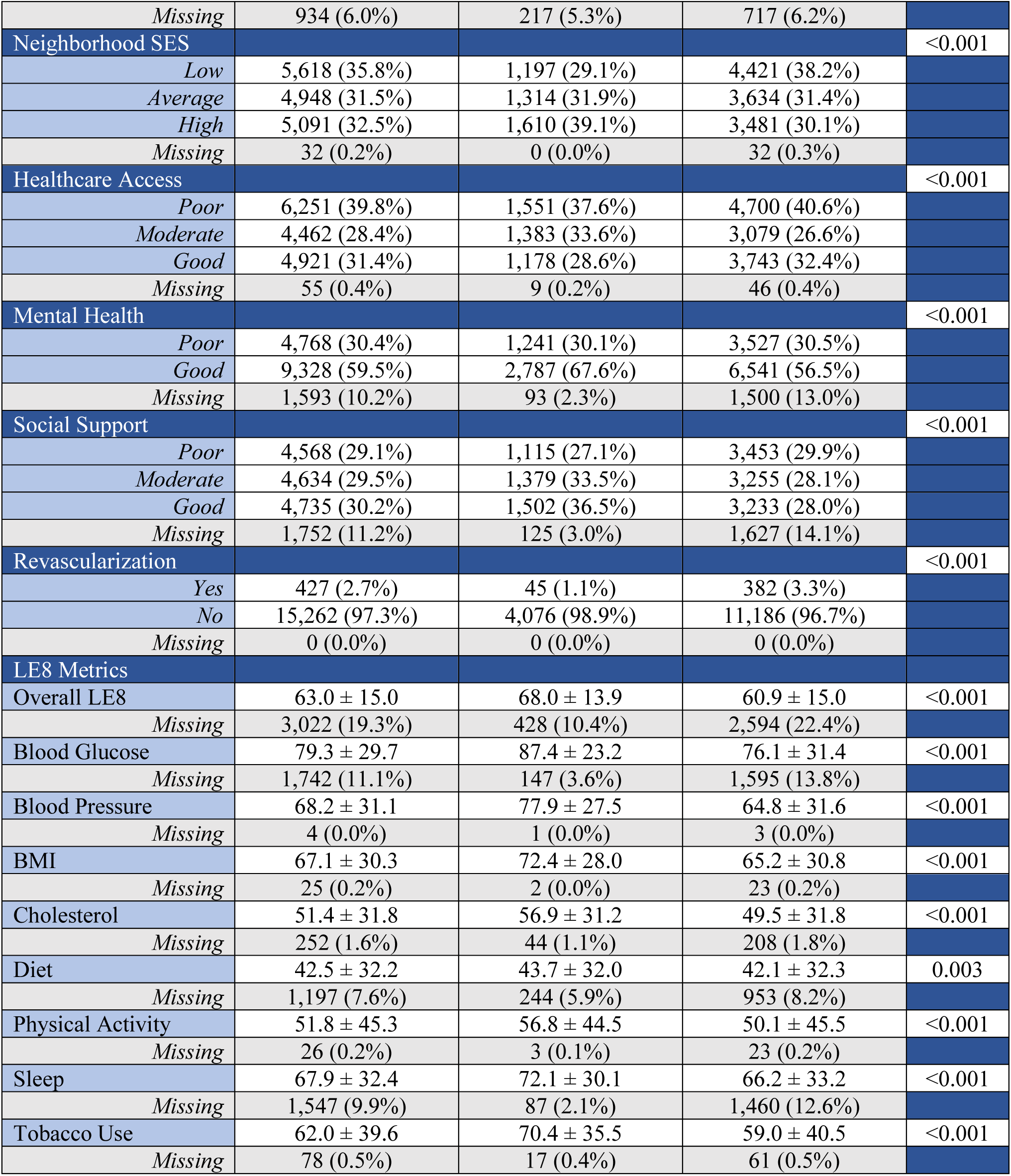
Demographics, social determinants of health, and LE8 metrics by trajectory analysis inclusion.

Nearly 84% of our study sample had complete covariate data at baseline. Covariate missingness for included participants ranged from 0.0% to 10.4%, with overall LE8 and individual SES missing the most data (Table 1). Missing data was determined to be missing at random; therefore, we built stabilized IPWs for missingness to account for missing data in our trajectory analysis. We modeled the bivariate association between each covariate and having complete baseline covariate data in our study using logistic regression to determine which variables to include in our IPWs. Healthcare access, individual SES, neighborhood SES, overall LE8, race-center, and sex were associated with missingness. Lastly, we combined our selection and missingness weights into an overall IPW.

### Statistical Analysis

We used group-based trajectory modeling (GBTM) to identify distinct trajectories of ABI values across ARIC Visits 1-7 (from 1987 to 2019). GBTM is rooted in the assumption that a population can be divided into distinct subgroups, each following a unique trajectory over time.^21^ First, we fit unconditional trajectory models with a censored normal distribution to identify a plausible number of trajectory groups using age as the timescale. To make this determination, we explored models with a varying number of groups, all set to a quadratic shape,^22^ and considered model fit statistics like Bayesian Information Criteria (BIC; lower values indicate better fit), latent class sizes (<5% is considered unstable), and *a priori* knowledge about ABIs and PAD. Second, we incorporated the overall IPW and covariates suspected to influence group membership (sex, race-center, history of CV disease, access to care, individual SES, neighborhood SES, mental health, and social support) into our models. To determine the ideal number and shape of each trajectory, we considered visual fit, latent class sizes, average posterior probabilities (>0.7 for each group is recommended), model fit statistics (e.g., BIC, Akaike Information Criteria, and log-likelihood), and overall interpretability.

Next, we compared weighted demographics, social determinants of health, and LE8 metrics across our final trajectory groups. We used chi-squared tests to compare weighted proportions, analysis of variance tests to compare normally distributed weighted means, and Kruskal-Wallis tests to compare non-normally distributed weighted means. In addition to our overall trajectory analysis, we replicated the previous steps within subgroups of sex (male and female) and race (Black and White) and conducted a sensitivity analysis where we included all limbs with ≥2 ABI measurements over the study period, expanding our original criteria of ≥3 ABI measurements. We performed all statistical analyses in SAS 9.4 (Cary, NC) and used PROC TRAJ for our GBTM analyses.

## Results

We identified 4,121 participants with three or more ABI measurements on at least one limb over the study period. Included participants had a baseline average age of 51.4 ± 4.9 years, were 57.3% female, 22.2% Black, and had an average overall LE8 score of 68.0 ± 13.9 points (Table 1). Most of the included limbs had three ABI measurements over the study period (73.5%), while 25.4% and 1.2% had four and five measurements, respectively.

### Trajectory model selection

Supplemental Table 3 outlines the various criteria used to evaluate each trajectory model. We prioritized visual fit, BIC, and substantive knowledge and found that models 3a and 4c were the best 3-and 4-group models, respectively. Model 3a had three linear trajectories, model 4c had three linear trajectories and one quadratic trajectory, and both models had low BICs and good visual fit. Additionally, both models displayed two steady linear trajectories within the normal ABI range; however, the 3-group model had one declining linear trajectory containing 8.3% of the weighted sample, while the 4-group model had two trajectories that declined over time, one quadratic and one linear, and contained 9.1% and 3.4% of the weighted sample, respectively. Substantively, both models were plausible, but the 4-group model had a group size <5%. Therefore, we chose model 3a as our final trajectory model and further explored the three distinct trajectories, which we named “low-normal,” “high-normal,” and “declining” (Figure 1).

**Figure 1.**
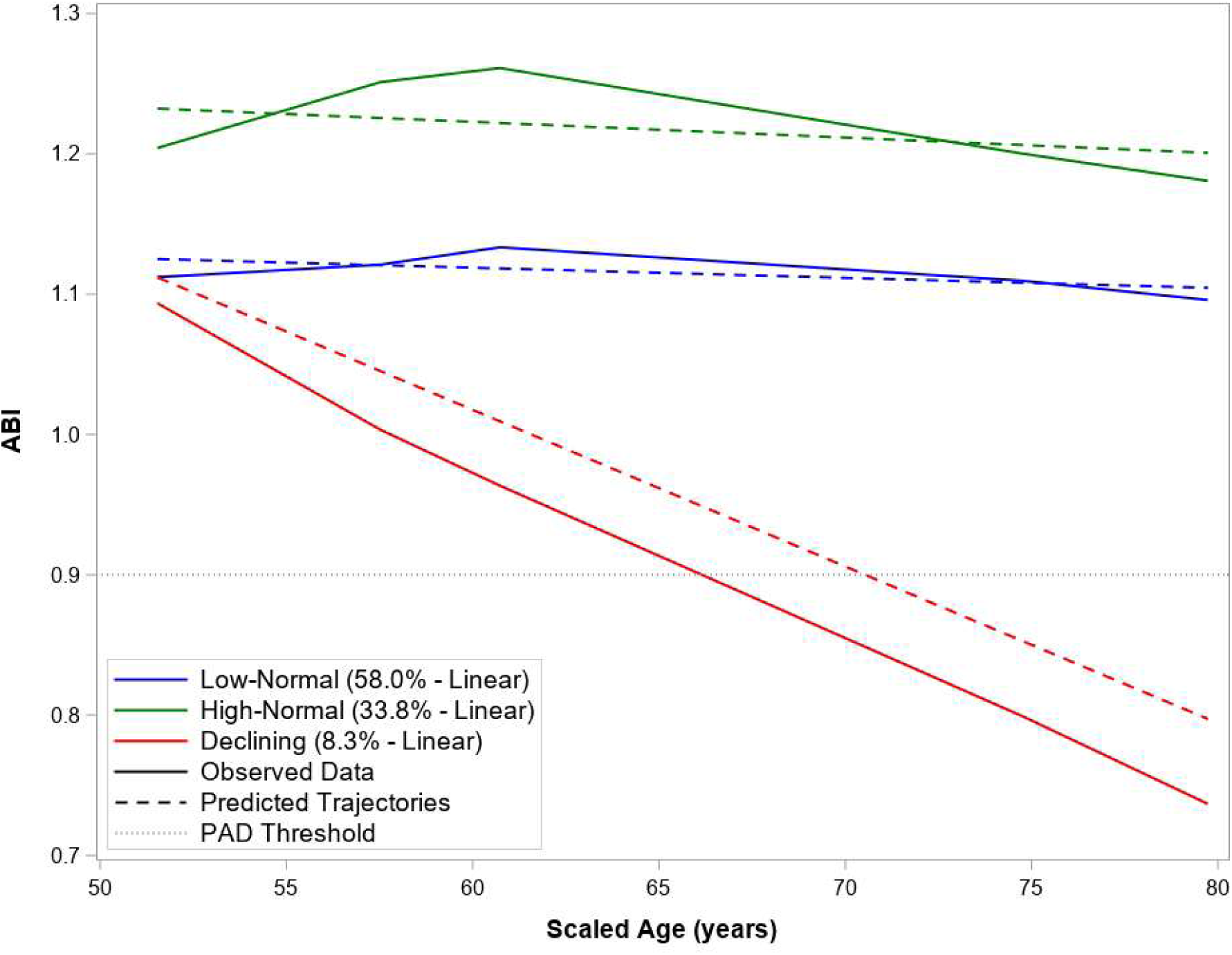
ABI Trajectories from Mid-to Late-Life, Best-Fitting 3-Group Model (Model 3a)

### Trajectory group demographics, social determinants of health, and LE8 metrics

The three trajectories in model 3a differed significantly by demographic factors, social determinants of health, and LE8 metrics (Table 2). The low-normal group contained the majority of the weighted sample (58.0%); while the high-normal and declining groups contained 33.8% and 8.3% of weighted participants, respectively. Baseline age did not differ significantly between trajectory groups. The low-normal group was 87.6% female, the high-normal group was primarily male (91.2%), and the declining group had a similar sex distribution to the overall sample. Participants in the declining group were more likely to be Black (41.3%), while those in the high-normal group were predominantly White (91.1%). Lastly, the declining group was more likely to have a history of CV disease, low individual and neighborhood SES, poor healthcare access, and poor social support.

**Table 2.**
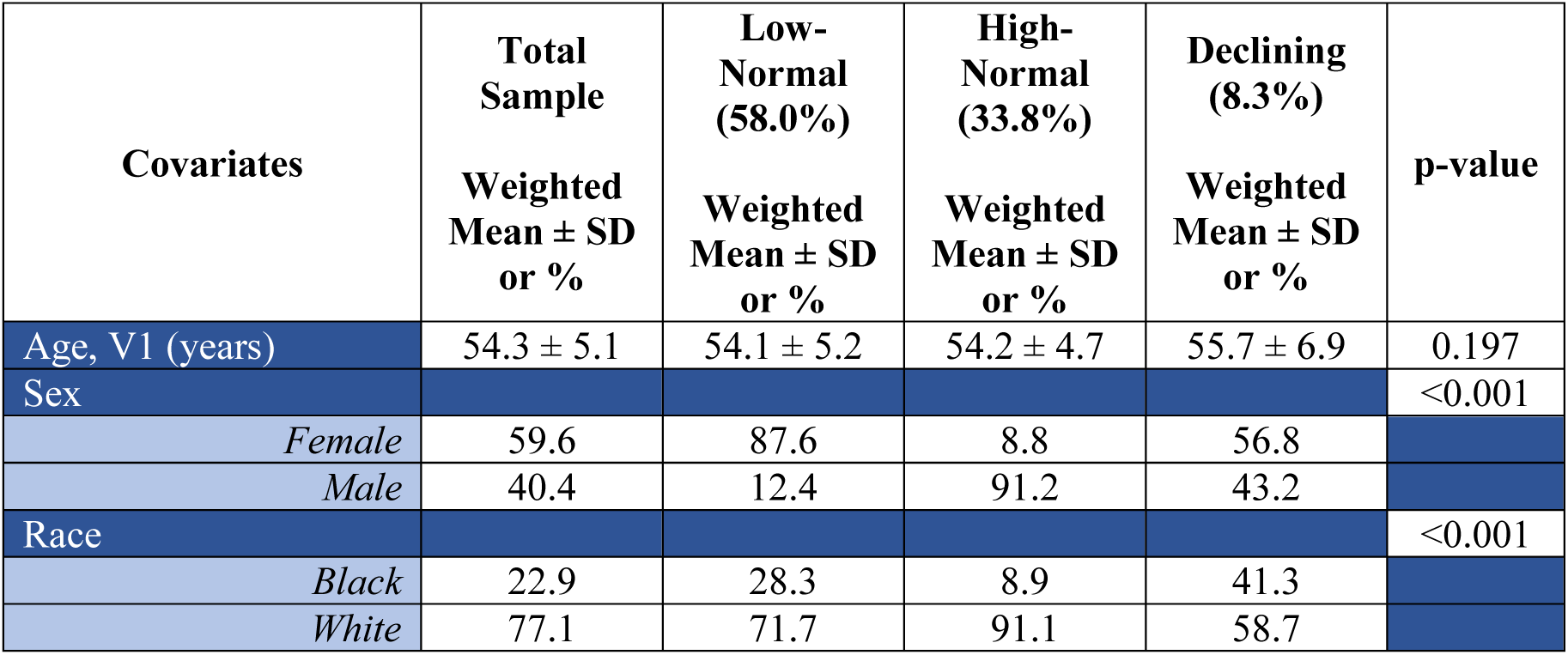
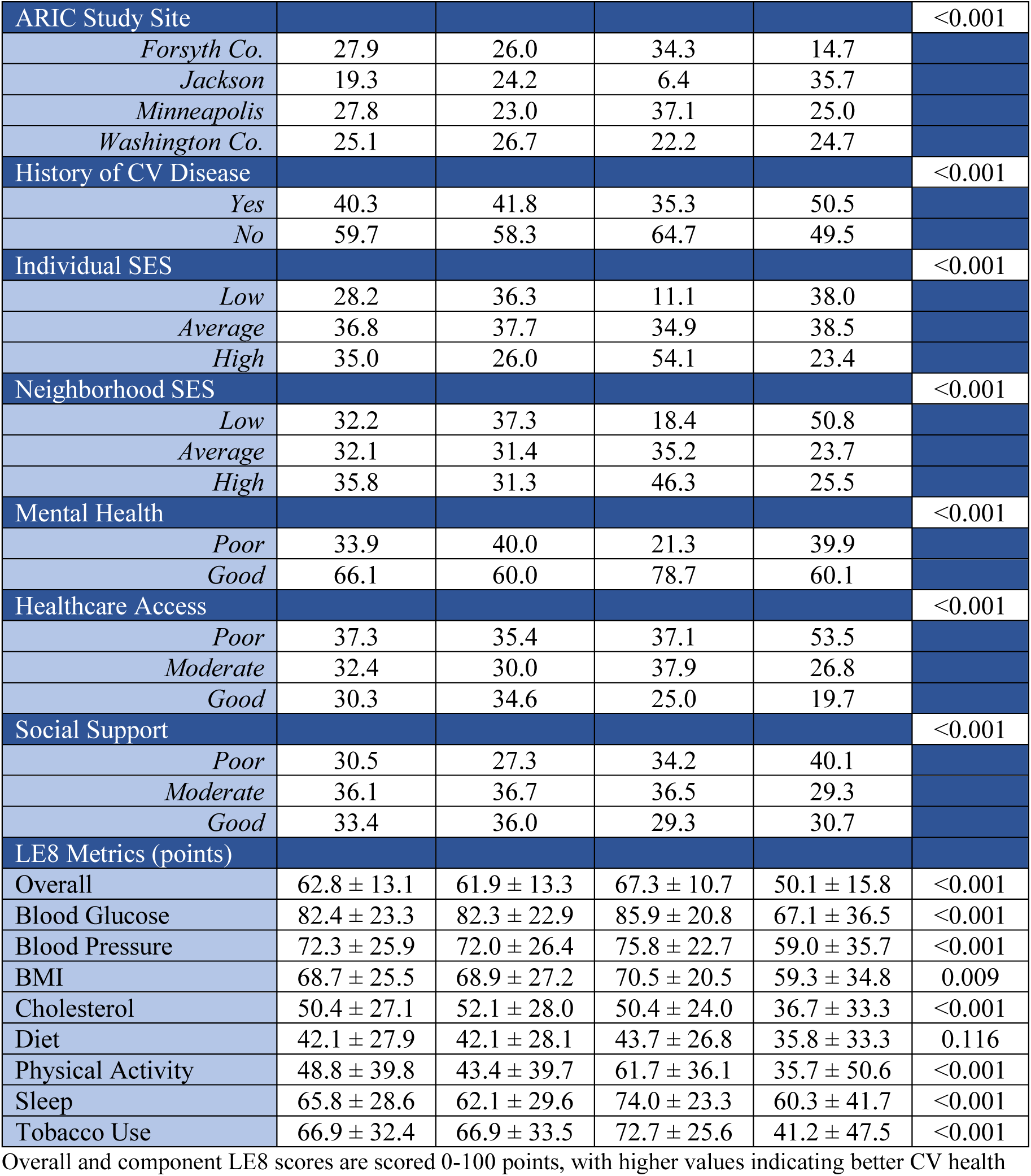
Weighted demographics, social determinants of health, and LE8 metrics for each trajectory group (Model 3a)

The total sample had a weighted average overall LE8 score of 62.8 ± 13.1 points (Table 2). In comparison, the high-normal group had a higher overall LE8 score (67.3 ± 10.7 points), while the declining group had a substantially lower overall LE8 scores (50.1 ± 15.8 points). Blood glucose had the highest average component score at 82.4 ± 23.3 points and diet had the lowest average score at 42.1 ± 27.9 points. When evaluating component LE8 scores within each trajectory group, some trends emerged. Overall, participants in the high-normal group had the highest component LE8 scores, while participants in the declining group had the lowest scores. Notably, four LE8 component scores (cholesterol, diet, physical activity, and tobacco use) fell within the poor CV health range for the declining group.

Compared to the high-normal group, overall LE8 scores in the low-normal and declining groups were 5.5 and 17.2 points lower, respectively (Figure 2). Five component LE8 scores were significantly lower in the declining group compared to the low-normal group, with tobacco use displaying the biggest difference (-25.7 points). All component LE8 metrics were significantly lower in the declining group compared to the high-normal group with one metric being 0-10 points lower, five metrics being 10-20 points lower, and two metrics (physical activity and tobacco use) being 20+ points lower.

**Figure 2.**
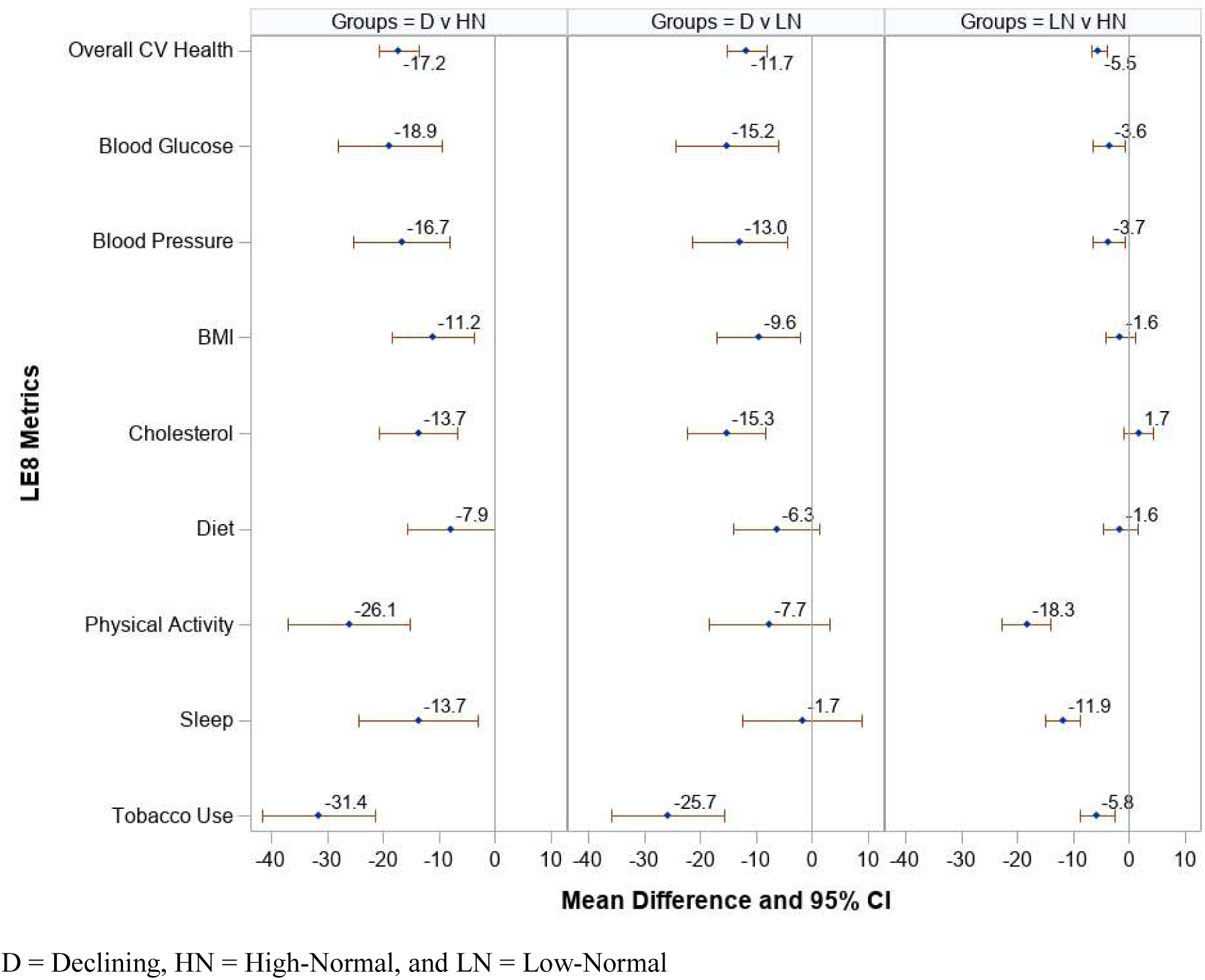
Weighted mean differences in LE8 scores between trajectory groups. D = Declining, HN = High-Normal, and LN = Low-Normal

### Subgroup and sensitivity analysis

For our subgroup analyses, trajectories for White, Black, and male participants displayed three distinct trajectories, two trajectories within the normal ABI range and one declining trajectory, similar to what we observed for the overall trajectories (Figure 3). For female participants, our 3-group trajectory analysis resulted in a group size <5%; therefore, we opted for a 2-group model with one steady, normal trajectory and one declining trajectory. Males had higher steady ABI trajectories than their female counterparts and more females experienced ABI decline compared to males (9.3% vs 5.3%). Additionally, among participants who experienced ABI decline, female participants showed a later onset of decline (around age 60 vs age 50), crossed the PAD threshold at an older age (74 vs 68 years), and had a smaller overall decline by age 80 (ABI = 0.81 vs 0.74) compared to male participants. When comparing trajectories among Black and White participants, 8.3% of Black participants experienced a decline in ABI compared to 7.6% of White participants. The trajectory of ABI decline differed by race: among Black participants, decline began sharply, likely before age 50, while decline for White participants began later, around age 60. Furthermore, among participants who experienced ABI decline, Black participants crossed the PAD threshold earlier, around age 60, compared to age 75 for White participants. By age 80, Black participants’ ABI had dropped further, to 0.70, compared to 0.84 for White participants. Lastly, although our sensitivity analysis (≥2 ABI measurements) increased the overall sample size to 8,658 participants, the results were not substantively different than the main results (Supplemental Figure 1).

**Figure 3.**
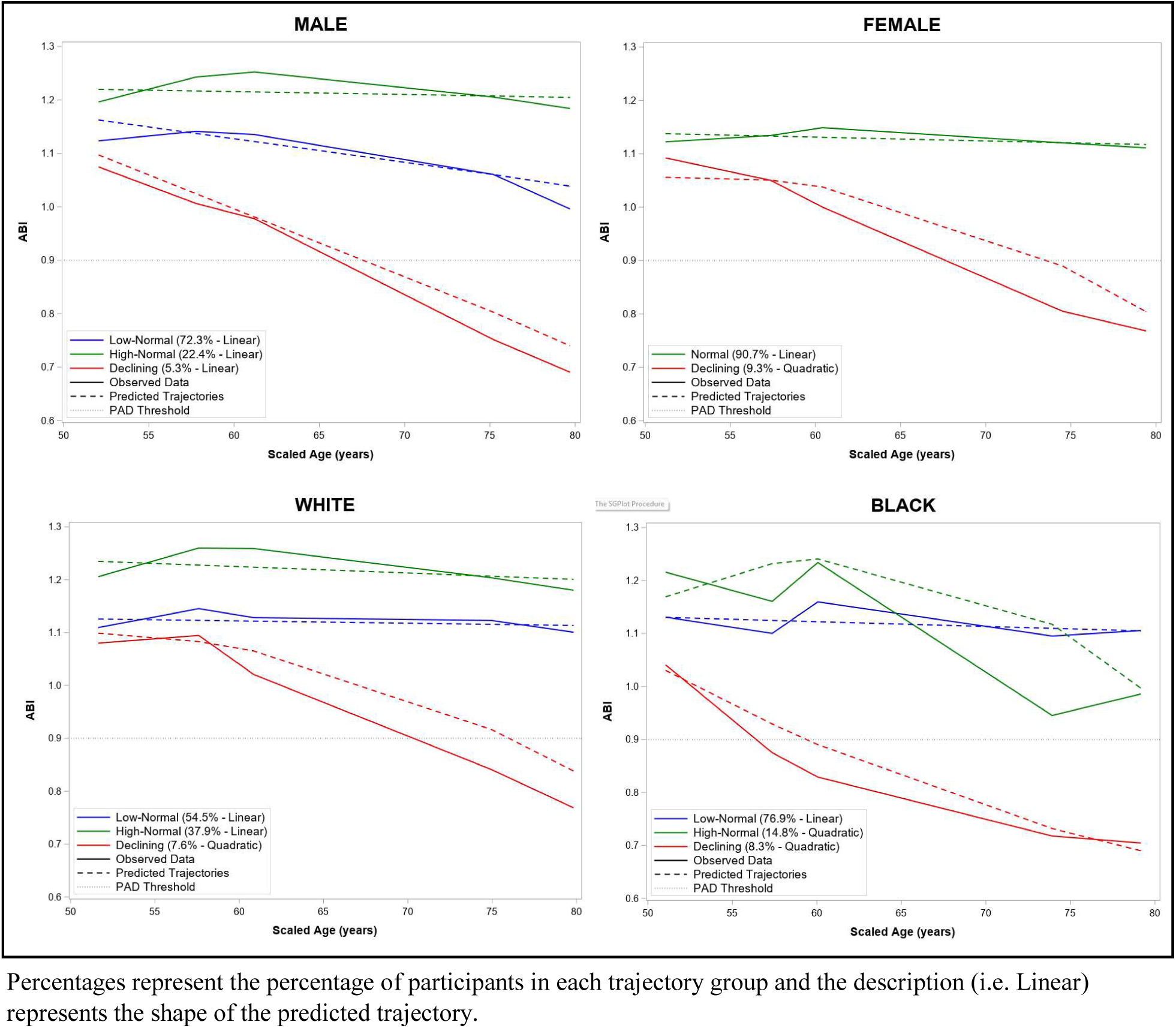
Subgroup ABI trajectories for sex and race. Percentages represent the percentage of participants in each trajectory group and the description (i.e. Linear) represents the shape of the predicted trajectory.

## Discussion

Traditionally, the ABI test is not offered to asymptomatic individuals, even if they have common PAD risk factors; consequently, the natural progression of ABI values from mid-to late-life remains poorly understood. Our overall trajectory analysis identified three distinct ABI trajectories throughout adulthood that were strongly associated with a various demographic, social, and CV health factors. To our knowledge, no prior studies have performed formal ABI trajectory modeling; however, our findings align with and extend previous longitudinal analyses of ABI values in asymptomatic populations. In one cohort of 76 Emory Healthcare patients, a meaningful ABI decline of 0.1 occurred over 8.3 years, with an average annual change of -0.012.^13^ This is comparable to the rate of decline observed in our declining group. Another Japanese cohort of 1,117 participants aged 18-76 years found ABI values increased on average up to age 50, providing insight into ABI behavior *prior* to the age range emphasized in our study.^14^

Although we identified three plausible ABI trajectories that are supported by prior literature, additional trajectories may exist. Specifically, a small portion of the population (∼2%) has arterial calcification resulting in ABI values >1.4.^23^ We did not observe a distinct trajectory in this range, but this was likely due to the small sample size of ABI values above 1.4. Furthermore, we uncovered a single declining trajectory, but it is possible that additional routes of decline exist. A large, longitudinal study focusing on individuals with ABI values outside of the normal range will be necessary to further explore possible ABI trajectories.

Participants with declining ABIs had significantly worst CV health compared to participants with stable ABI values. Low ABI values, and thus PAD, are often the result of uncontrolled modifiable risk factors, like diabetes, hypertension, and smoking.^24^ One study of 155,722 participants without a history of CV disease estimated that ∼70% of CV events and death were attributable to modifiable risk factors.^25^ To better define ideal CV health, the American Heart Association developed Life’s Simple 7 (LS7) in 2010 and expanded this framework in 2022 to LE8 with the addition of sleep health.^19,26^ Hundreds of papers have since used these frameworks to assess the effect of overall CV health on multiple CV diseases and outcomes, with a handful focusing on PAD. Two studies, one among 12,865 ARIC participants aged 45-64 years at baseline and another among 5,529 participants aged 45-84 years at baseline from the Multi-Ethnic Study of Atherosclerosis (MESA) study, found that higher LS7 scores were associated with a lower risk of PAD.^27,28^ Another study assessing the association between LE8 and PAD in the 1999-2004 National Health and Nutrition Examination Survey (NHANES) similarly concluded that participants aged 20 years or older with high CV health had a lower prevalence of PAD.^29^

Five of the eight component LE8 scores (blood glucose, blood pressure, BMI, cholesterol, and tobacco use) were significantly lower in the participants experiencing ABI decline compared to those who did not. Numerous studies have found strong associations between these risk factors and ABI decline and PAD.^30–32^ In particular, a study of older adults in the Cardiovascular Health Study found that ABI decline was independently associated with current cigarette use, hypertension, diabetes, and higher low-density lipoprotein cholesterol.^12^ We also observed that participants with declining ABI values were more likely to have a history of CV disease. This is supported by an evaluation of primary care in the US where 50% of patients with PAD had co-occurring CV disease.^1,33,34^ Together, these findings reinforce the central role of modifiable risk factors and overall CV health in ABI decline. Middle-aged individuals with poor overall CV health, multiple modifiable risk factors, especially tobacco use, diabetes, hypertension, and high cholesterol, or a history of CV disease should be considered high-risk for incident PAD and may benefit from screening with the ABI test.

Subgroup analyses revealed important sex and racial differences in baseline ABI values, group sizes, route of ABI decline, and level of decline. Women had lower steady ABI trajectories than men, consistent with a cross-sectional study of 1,775 healthy MESA participants that found females, regardless of race/ethnicity, had lower baseline ABI values compared to their male counterparts.^35^ The authors attributed this to biological differences such as height and arterial geometry. Additionally, a larger proportion of women experienced declining ABI values, even after incorporating various demographic, social, and CV-health-related factors into our trajectory models. This aligns with a systematic review of 118 articles that found the prevalence of PAD to be slightly higher in women compared to men in high income countries (7.81% vs 6.60% at 55-59 years) and another study pooling data from six US community-based cohorts (5.0% of men vs 5.9% of women aged 40+).^36,37^ Lastly, we found that, compared to men, women had a delayed onset of ABI decline by roughly 10 years. A review of PAD in women discussed how women present with PAD 10 to 20 years later than men, on average, and noted this often corresponds with the onset of menopause, suggesting estrogen may be protective against ABI decline and PAD.^38^ These findings suggest that while high-risk individuals of both sexes may benefit from ABI screening, the optimal timing of such screening may differ by sex.

Black participants were more likely than White participants to experience ABI decline with earlier onset, faster progression, and greater severity. This parallels cohort data demonstrating higher PAD prevalence among non-Hispanic Black individuals (Range: 7.2%-21.5%) compared to non-Hispanic White individuals (Range: 3.6%-11.9%).^12,39–42^ Current evidence suggests these disparities are largely driven by social determinants of health.^43^ One review summarized that Black individuals are more likely to have multiple CV risk factors, like diabetes, hypertension, and obesity, and comorbid CV disease.^44^ This review also discussed that Black individuals have a higher prevalence of asymptomatic PAD, present at younger ages with more severe disease, and experience worse outcomes compared to their White counterparts. Therefore, earlier ABI screening in high-risk Black populations may be warranted.

The USPSTF currently recommends several sex-specific screenings based on differential risk, including prostate and cervical cancer screening.^45,46^ Additionally, breast cancer and osteoporosis screenings are emphasized for women, whereas abdominal aortic aneurysm screening is primarily recommended for men, although both sexes can develop these conditions.^47–49^ In contrast, race-specific screening guidelines are uncommon despite well-documented disparities. This is largely due to insufficient race-specific evidence although emerging data highlights the potential value of tailored screening approaches by race. For example, a 10-year nationwide study of more than 400,000 breast cancer deaths found that Black women had a substantially higher risk of death from early-onset breast cancer compared to White women.^50^ Additionally, they found that breast cancer screening for Black women should begin up to 8 years earlier than the recommended starting age of 50. Sex-and race-specific ABI screening strategies may similarly improve early PAD detection, though additional research on the benefits and harms of screening for PAD with the ABI test is required.

To our knowledge, this is the first study to conduct a formal trajectory analysis of ABI values from mid-to late-life and assess the association between distinct ABI trajectories and CV health, measured using the LE8 framework. We utilized data from the ARIC study, a large, longitudinal cohort study that collected the variables necessary to calculate baseline LE8 scores and repeat ABI values over 30-years beginning in mid-life. No other cohort has collected multiple ABI values on healthy middle-aged participants over a period that long. In addition to these strengths, we also encountered a few limitations. Firstly, a large number of participants were excluded from this study for having <3 ABI measurements on at least one limb over the study-period and excluded participants reported more disparities and had worse CV health compared to included participants. We were able to mitigate this bias by incorporating stabilized IPWs to make our results more representative of the overall population, although some bias may persist. Secondly, we were only able to calculate LE8 scores at baseline, so we were unable to analyze how changing LE8 scores may have influenced our ABI trajectories. Lastly, four of our covariates were collected at ARIC Visit 2 and used as a proxy for ARIC Visit 1 and a handful of variables were self-reported measures (Supplemental Table 1), which may have introduced some measurement error into our analysis. Generalizing these results to the broader populations should be done with caution, as ARIC is representative only of its four original study communities.

## Conclusions

We identified three distinct ABI trajectories from mid-to late-life, including one characterized by progressive decline into the PAD range. Participants who experienced ABI decline exhibited substantially poorer overall CV health and higher burdens of modifiable risk factors at baseline. Targeted ABI screening in middle-aged adults with poor CV health, potentially with sex-and race-informed timing, may improve early PAD detection and intervention. Further research is needed to confirm these findings and evaluate the benefits and harms of screening for PAD with the ABI test.

## Data Availability

This project utilized data from the Atherosclerosis Risk in Communities (ARIC) Study and required an accepted project proposal to access the data. Therefore, the data will not be made publicly available.

## Acknowledgments

The Atherosclerosis Risk in Communities study has been funded in whole or in part with Federal funds from the National Heart, Lung, and Blood Institute, National Institutes of Health, Department of Health and Human Services, under contract numbers (HHSN268201700001I, HHSN268201700002I, HHSN268201700003I, HHSN268201700004I, and HHSN268201700005I). The authors thank the staff and participants of the ARIC study for their important contributions.

---

**Supplemental Table 1.** Original Variables and Final Coding Descriptions for All Variables.

| Important Variables | Original Questions and Responses | Final Coding |
| --- | --- | --- |
| **Limb-Related Variables** |  |  |
| Ankle Brachial Index (ABI) |  |  |
| *Visit 1* | Continuous (range: 0.37-1.80) | Same |
| *Visit 3* | Continuous (range: 0.29-1.74) | Same |
| *Visit 4* | Continuous (range: 0.41-1.97) | Same |
| *Visit 5, Left* | Continuous (range: 0.41-1.94) | Same |
| *Visit 5, Right* | Continuous (range: 0.36-1.71) | Same |
| *Visit 7, Left* | Continuous (range: 0.41-1.81) | Same |
| *Visit 7, Right* | Continuous (range: 0.23-1.71) | Same |
| ABI Limb |  |  |
| *Visit 1* | Right or left | Same |
| *Visit 3* | Right or left | Same |
| *Visit 4* | Right or left | Same |
| **Demographics** |  |  |
| Age |  |  |
| *Visit 1* | Continuous (range: 44-66) | Same |
| *Visit 3* | Continuous (range: 49-73) | Same |
| *Visit 4* | Continuous (range: 52-75) | Same |
| *Visit 5* | Continuous (range: 67-90) | Same |
| *Visit 7* | Continuous (range: 75-98) | Same |
| Sex | Male or female | Same |
| Race | Asian, Black, American Indian or Alaskan Indian, or White | Non-White and non-Black participants were excluded from this analysis. Due to collinearity between the race and center variables, we created a race-center variable with eight groups: BF, BJ, BM, BW, WF, WJ, WM, WW |
| ARIC Field Center | Forsyth County, NC; Jackson, MS; Minneapolis; Washington County, MD | Due to collinearity between the race and center variables, we created a race-center variable with eight groups: BF, BJ, BM, BW, WF, WJ, WM, WW |
| **Covariates** |  |  |
| Access to Care (Visit 1) | (1) Time in months since last saw a doctor (Responses: 6 months or less, 6 months to 1 year, 1-2 years, >2 years) | (1) 6 months or less = 1 point; 6 months to 1 year = 0.66 points; 1-2 years = 0.33 points; |
|  | (2) How often do you have a routine physical exam, not for a particular illness, but for a general checkup?<br>(Responses: At least once a year, at least once every five years, less than once every five years, do not have routine physical examinations)<br>(3) Has health insurance (yes/no) | >2 years = 0 points.<br>(2) At least once a year = 1 point; At least once every five years = 0.66 points; Less than once every five years = 0.33 points; Do not have routine physical examinations = 0 points.<br>(3) Yes = 1 point; No = 0 points.<br>We combined points from all three access to care questions and created tertile groups (poor, moderate, good).<br>Higher scores indicate better access to care. |
| History of CVD (Visit 1) | (1) Has a doctor ever said you had a heart attack?<br>(2) Has a doctor ever said you had a stroke?<br>(3) Doctor diagnosis of chest pain.<br>(4) Have you ever had a heart attack for which you were hospitalized one week or more?<br>(5) Were you hospitalized for this problem in your legs?<br>(6) Have you ever had to sleep on 2 or more pillows to help you breathe?<br>(7) Breathing trouble ever wake you?<br>(8) Have you ever had surgery on your heart, or on the arteries of your neck or legs, excluding surgery for varicose veins?<br>(9) Balloon angioplasty?<br>(10) Carotid bruits?<br>(11) Rales?<br>(12) Systolic murmur?<br>(13) Diastolic murmur?<br>(14) Arrhythmia? | If the answer was “yes” to any of these questions then history of CVD was “yes.” Otherwise, history of CVD was “no.” |
| Individual socioeconomic status (SES) (Visit 1) | (1) Highest education completed, number of grades (range: 0-23)<br>(2) Combined family income:<br>1 = Under \$5,000<br>2 = \$5,000-\$7,999<br>3 = \$8,000-\$11,999<br>4 = \$12,000-\$15,999<br>5 = \$16,000-\$24,999<br>6 = \$25,000-\$34,999<br>7 = \$35,999-\$49,999<br>8 = Over \$50,000 | We created z-scores for both education and income, combined them into one overall z-score, then created tertile groups (low, average, high).<br>Higher scores indicate better individual SES. |
| Mental Health (Visit 2) | (1) Do you believe that you have come to a “dead end”?<br>(2) Have you ever experienced a feeling of hopelessness recently?<br>(3) Do little things irritate you more lately than they used to?<br>(4) Do you feel you want to give up trying?<br>(5) Would you want to be dead at times?<br>(6) Do you feel rejected?<br>(7) Do you feel like crying sometimes?<br>Responses to the following questions were “yes,” “don’t know,” or “no.” | Yes = 2 points, Don’t Know = 1 point, No = 0 points.<br>We combined points from all seven questions and created a binary variable (good/poor) with a median split (median = 2 points).<br>Fewer points indicate better mental health. |
| Neighborhood SES (Visit 1) | Neighborhood SES summary z-score | We created tertile groups from the summary z-scores (low, average, high).<br>Higher scores indicate better neighborhood SES. |
| Overall LE8 | -- | Overall LE8 scores are the average of the eight component LE8 scores (described below), calculated as the sum of all eight component scores divided by eight. These scores are a continuous, ranging from 0-100 points. |
|  |  | Higher points indicate better overall CV health. |
| *Blood Glucose*<br>*(ARIC Visit 2)* | (1) Hemoglobin A1c (HbA1c; range: 3.5-18.2)<br>(2) Ever diagnosed with diabetes (yes/no) | LE8 points for blood glucose were assigned as follows:<br>100 = No history of diabetes and HbA1c <5.7<br>60 = No diabetes and HbA1c 5.7-6.4)<br>40 = Diabetes with HbA1c <7.0<br>30 = Diabetes with HbA1c 7.0-7.9<br>20 = Diabetes with HbA1c 8.0-8.9<br>10 = Diabetes with HbA1c 9.0-9.9<br>0 = Diabetes with HbA1c ≥10.0<br>Higher points indicate better blood glucose LE8 scores. |
| *Blood Pressure*<br>*(ARIC Visit 1)* | (1) Average of the 2<sup>nd</sup> and 3<sup>rd</sup> of three systolic BP measurements (range: 61.0 – 246.0 mmHg)<br>(2) Average of the 2<sup>nd</sup> and 3<sup>rd</sup> of three diastolic BP measurements (range: 0.0 – 144.0 mmHg)<br>(3) Hypertension lowering medication within past 2 weeks (yes/no) | LE8 points for blood pressure were assigned as follows:<br>100 = <120 / <80 mmHg<br>75 = 120-129 / <80 mmHg<br>50 = 130-139 or 80-89 mmHg<br>25 = 140-159 or 90-99 mmHg<br>0 = ≥160 or ≥100 mmHg<br>(Subtract 20 points if treated level) |
|  |  | Higher points indicate better blood pressure LE8 scores. |
| *BMI (ARIC Visit 1)* | Continuous BMI in kg/m<sup>2</sup><br>(range: 14.2-65.9 kg/m<sup>2</sup>) | LE8 points for BMI were assigned as follows:<br>100 = <25 kg/m<sup>2</sup><br>70 = 25.0-<30.0 kg/m<sup>2</sup><br>30 = 30.0-<35.0 kg/m<sup>2</sup><br>15 = 35.0-<40.0 kg/m<sup>2</sup><br>0 = ≥40.0 kg/m<sup>2</sup><br>Higher points indicate better BMI LE8 scores. |
| *Cholesterol (ARIC Visit 1)* | (1) Plasma total cholesterol (TC; range: 48.0-594.0 mg/dL)<br>(2) High Density Lipoprotein (HDL/good cholesterol; range: 9.6-163.0 mg/dL)<br>(3) Has taken cholesterol lowering medication within the past 2 weeks (yes/no) | Plasma total cholesterol and HDL cholesterol used to calculate non-HDL cholesterol (TC – HDL = non-HDL). LE8 points for non-HDL were assigned as follows:<br><130 mg/dL = 100 points<br>130-159 mg/dL = 60 points<br>160-189 mg/dL = 40 points<br>190-219 mg/dL = 20 points<br>≥220 mg/dL = 0 points<br>(If drug-treated level, subtract 20 points)<br>Higher points indicate better cholesterol LE8 scores. |
| *Diet (ARIC Visit 1)* | (1) <u>Dairy</u>: skim or low-fat milk; yogurt; cottage cheese<br>(2) <u>Fruit</u>: apples or pears; oranges; fruit juice; peaches, apricots, plums; bananas; other fruits | Scores for all foods within each category (e.g. dairy, fruit, etc.) were combined, then we created quintile groups for each category. Lastly, we combined all |
|  | (3) <u>Vegetables</u>: string or green beans; broccoli; cabbage, cauliflower, brussels sprouts; carrots; corn; spinach, collards, greens; dark yellow squash; sweet potatoes; tomatoes<br>(4) <u>Nuts/legumes</u>: peas or lima beans; beans or lentils; peanut butter; nuts<br>(5) <u>Whole grains</u>: cooked cereals, dark or grain breads<br>(6) <u>Sodium</u>: How often is salt or salt-containing seasoning added to your food in cooking? How many shakes of salt do you add to your food at the table every day?<br>(7) <u>Red and processed meats</u>: hamburger; hot dogs; processed meats; bacon; beef, pork, lamb as side dish; beef pork, lamb as main dish<br>(8) <u>Sugar-sweetened beverages</u>: low calorie soft drinks, regular soft drinks, fruit-flavored punch<br>Responses for the number of servings consumed for each food (e.g. skim milk, regular soft drinks, etc.) includes:<br>• Never (where applicable) = 0 points<br>• Almost never = 1 point<br>• 1-3 times per month = 2 points<br>• 1 per week = 3 points<br>• 2-4 per week = 4 points<br>• 5-6 per week = 5 points<br>• 1 per day = 6 points<br>• 2-3 per day = 7 points<br>• 4-6 per day = 8 points<br>• >6 per day = 9 points | category scores and assigned diet LE8 points as follows:<br>$\geq 95^{\text{th}}$ percentile (top/ideal) = 100 points<br>$75^{\text{th}}-94^{\text{th}}$ percentile = 80 points<br>$50^{\text{th}}-74^{\text{th}}$ percentile = 50 points<br>$25^{\text{th}}-49^{\text{th}}$ percentile = 25 points<br>$1^{\text{st}}-24^{\text{th}}$ percentile (bottom/least ideal) = 0 points<br>Higher points indicate better diet LE8 scores. |
|  | Reverse coding for sodium, red and processed meats, and sugar-sweetened beverage foods. |  |
| *Physical Activity (ARIC Visit 1)* | Minutes of moderate to vigorous exercise per week | Physical activity LE8 points were assigned as follows:<br>≥150 mins = 100 points<br>120-149 mins = 90 points<br>90-119 mins = 80 points<br>60-89 mins = 60 points<br>30-59 mins = 40 points<br>1-29 mins = 20 points<br>0 mins = 0 points<br>Higher points indicate better physical activity LE8 scores. |
| *Sleep (ARIC Visit 2)* | (1) Do you often feel tired?<br>(2) Do you often have trouble falling asleep?<br>(3) Do you wake up repeatedly during the night?<br>Responses to the following questions were “yes,” “don’t know,” or “no.” | Yes = 2 points, Don’t Know = 1 point, No = 0 points. We combined points from all 3 questions and sleep LE8 points were assigned as follows:<br>0 = 100 points<br>1 = 90 points<br>2 = 70 points<br>3 = 60 points<br>4 = 40 points<br>5 = 20 points<br>6 = 0 points<br>Higher points indicate better sleep LE8 scores. |
| *Tobacco Use*<br>*(ARIC Visit 1)* | (1) Have you ever smoked cigarettes? (yes/no)<br>(2) Do you now smoke cigarettes? (yes/no)<br>(3) Number of years abstained from cigarettes. (range: 0-40 years)<br>(4) Number of hours a week around smokers. (range: 0-125 hrs/week) | Tobacco use LE8 points were assigned as follows:<br>Never smoker = 100 points<br>Former smoker, quit $\geq 3$ years, or non-smoker in the upper quartile of hours/week of exposure to second hand smoke = 75 points<br>Former smoker, quit 1-3 years = 50 points<br>Former smoker, quit <1 year = 25 points<br>Current smoker = 0 points<br>Higher points indicate better tobacco use LE8 scores. |
| Revascularization | ICD-9 Codes: 38.18, 39.25, 39.29, 39.50<br>CPT Codes: 04Cxxxx, 041xxxx, 047xxxx (882 codes) | If any code present, then revascularization = yes<br>Otherwise, revascularization = no |
| Date of Revascularization | If revascularization = yes, this variable represents when the revascularization occurred in the number of days since enrollment in the ARIC study. | Same |
| Social Support (Visit 2) | (1) Most of my friends are more interesting than I am.<br>(2) When I feel lonely, there are several people I can talk to.<br>(3) I often meet or talk with family or friends.<br>(4) I feel like I'm not always included by my circle of friends.<br>(5) There really is no one who can give me an objective view of how I'm handling my problems.<br>(6) If I were sick and needed someone (friend, family | Definitely true = 3 points, Probably true = 2 points, Probably false = 1 point, Definitely false = 0 points<br>Questions 1, 4, 5, 6, 9, 10, 11, 12, 14, 16 were reverse coded.<br>We combined points from all 16 questions and calculated tertiles to create a three-level categorical variable (low, average, high). |
|  | member, or acquaintance) to take me to the doctor, I would have trouble finding someone.<br>(7) If I were sick, I could easily find someone to help me with my daily chores.<br>(8) When I need suggestions on how to deal with a personal problem, I know someone I can turn to.<br>(9) I often don't get invited to do things with others.<br>(10) Most of my friends are more successful at making changes in their lives than I am.<br>(11) If I had to go out of town for a few weeks, it would be difficult to find someone who would look after my house or apartment (the plants, pets, garden, etc.).<br>(12) There is really no one I can trust to give me good financial advice.<br>(13) I am more satisfied with my life than most people are with theirs.<br>(14) It would be difficult to find someone who would lend me their car for a few hours.<br>(15) There is at least one person I know whose advice I really trust.<br>(16) I have a hard time keeping pace with my friends.<br>Responses are "definitely true," "probably true," "probably false," or "definitely false." | Higher points indicate better social support. |

**Supplemental Table 2.**
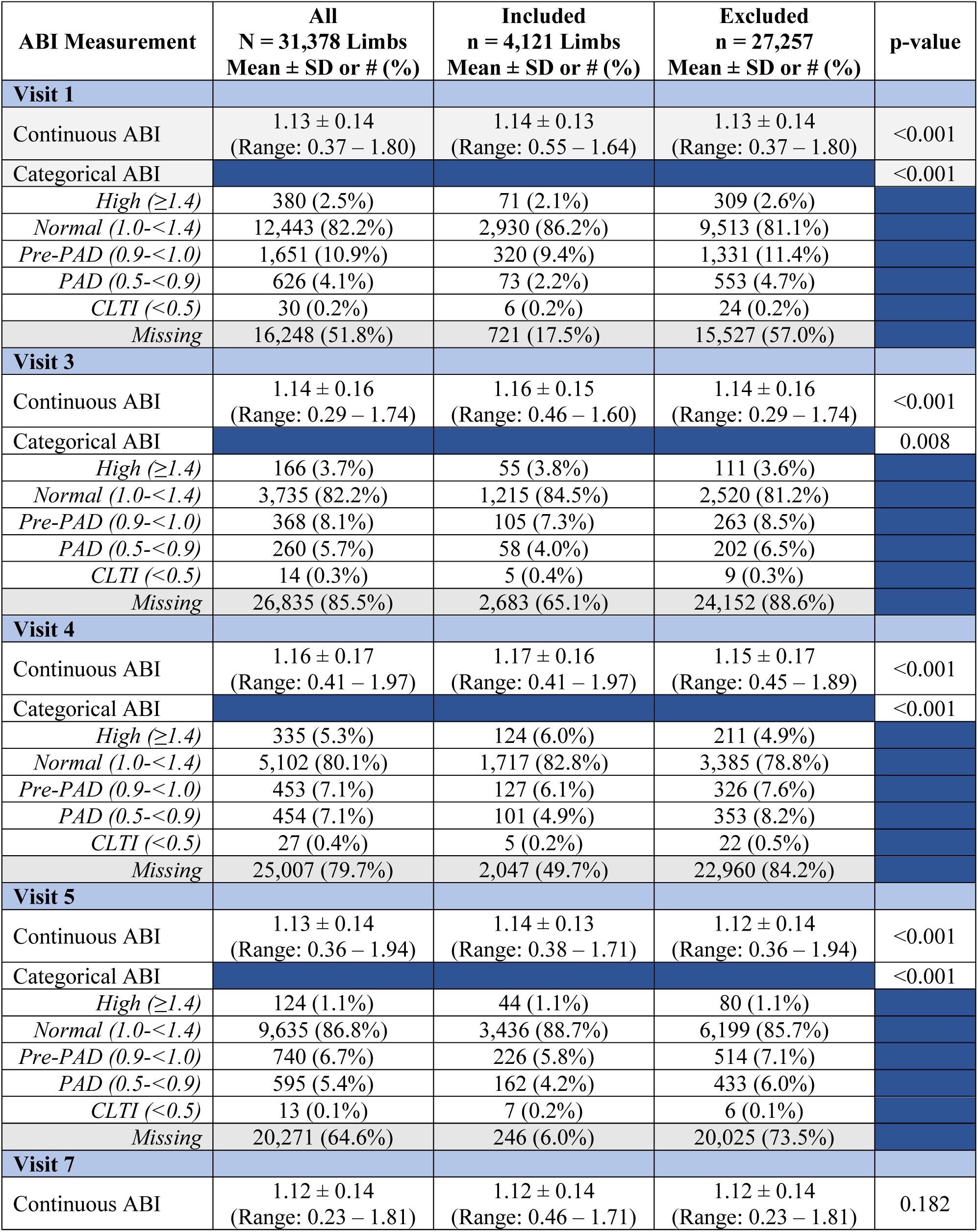
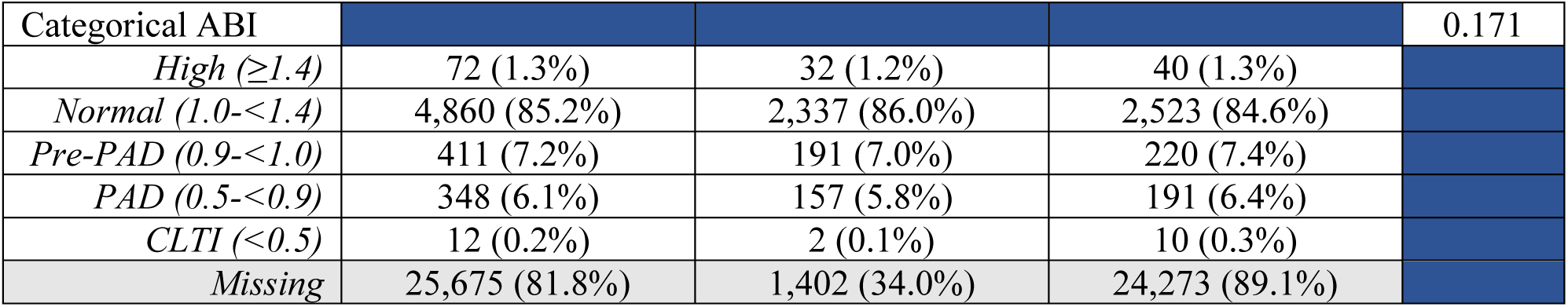
Continuous and categorical ABI values by visit and trajectory analysis inclusion.

**Supplemental Table 3.** GBTM Fit Statistics by Number of Groups and the Shape of Each Group.

| Model/<br># Groups | Shape | Group<br>% | Average<br>Posterior Prob. | BIC | AIC | Log<br>Likelihood | Visual<br>Fit |
| --- | --- | --- | --- | --- | --- | --- | --- |
| **Overall Trajectory Groups** |  |  |  |  |  |  |  |
| 3a* | Linear | 58.0 | 0.855 | 6101.62 | 6200.63 | 6227.63 | Good |
|  | Linear | 33.8 | 0.839 |  |  |  |  |
|  | Linear | 8.3 | 0.875 |  |  |  |  |
| 3b | Quadratic | 55.6 | 0.840 | 6233.11 | 6343.13 | 6373.13 | Okay |
|  | Quadratic | 35.5 | 0.842 |  |  |  |  |
|  | Quadratic | 8.8 | 0.879 |  |  |  |  |
| 3c | Linear | 57.6 | 0.850 | 6123.51 | 6226.19 | 6254.19 | Okay |
|  | Linear | 33.6 | 0.838 |  |  |  |  |
|  | Quadratic | 8.8 | 0.879 |  |  |  |  |
| 3d | Quadratic | 46.3 | 0.856 | 6190.84 | 6293.52 | 6321.52 | Good |
|  | Linear | 45.7 | 0.797 |  |  |  |  |
|  | Linear | 8.1 | 0.869 |  |  |  |  |
| 3e | Quadratic | 45.8 | 0.857 | 6210.69 | 6227.88 | 6346.03 | Okay |
|  | Linear | 45.6 | 0.790 |  |  |  |  |
|  | Quadratic | 8.6 | 0.859 |  |  |  |  |
| 4a | Linear | 55.7 | 0.848 | 6200.70 | 6343.72 | 6382.72 | Poor |
|  | Linear | 35.9 | 0.838 |  |  |  |  |
|  | Linear | 4.2 | 0.851 |  |  |  |  |
|  | Linear | 4.1 | 0.804 |  |  |  |  |
| 4b | Quadratic | 53.4 | 0.826 | 6402.17 | 6559.86 | 6602.86 | Okay |
|  | Quadratic | 35.3 | 0.837 |  |  |  |  |
|  | Quadratic | 7.4 | 0.795 |  |  |  |  |
|  | Quadratic | 4.0 | 0.813 |  |  |  |  |
| 4c* | Linear | 50.3 | 0.809 | 6261.61 | 6408.29 | 6448.29 | Good |
|  | Linear | 37.3 | 0.834 |  |  |  |  |
|  | Quadratic | 9.1 | 0.709 |  |  |  |  |
|  | Linear | 3.4 | 0.849 |  |  |  |  |
| 4d | Linear | 52.5 | 0.818 | 6280.75 | 6431.10 | 6472.10 | Good |
|  | Linear | 35.1 | 0.835 |  |  |  |  |
|  | Quadratic | 9.1 | 0.766 |  |  |  |  |
|  | Quadratic | 3.4 | 0.867 |  |  |  |  |
| 4e | Linear | 44.2 | 0.852 | 6371.21 | 6525.24 | 6567.24 | Okay |
|  | Quadratic | 44.1 | 0.782 |  |  |  |  |
|  | Quadratic | 8.5 | 0.776 |  |  |  |  |
|  | Quadratic | 3.2 | 0.855 |  |  |  |  |

| Male |  |  |  |  |  |  |  |
| --- | --- | --- | --- | --- | --- | --- | --- |
| M3a* | Linear | 72.3 | 0.920 | 2402.14 | 2482.58 | 2507.58 | Good |
|  | Linear | 22.4 | 0.725 |  |  |  |  |
|  | Linear | 5.3 | 0.868 |  |  |  |  |
| M3b | Quadratic | 65.3 | 0.885 | 2480.91 | 2571.01 | 2599.01 | Good |
|  | Quadratic | 27.3 | 0.726 |  |  |  |  |
|  | Quadratic | 7.4 | 0.897 |  |  |  |  |
| M3c | Linear | 50.5 | 0.819 | 2466.48 | 2550.15 | 2576.15 | Okay |
|  | Quadratic | 41.6 | 0.748 |  |  |  |  |
|  | Linear | 7.9 | 0.915 |  |  |  |  |
| M3d | Linear | 72.4 | 0.920 | 2403.34 | 2485.00 | 2511.00 | Good |
|  | Linear | 21.8 | 0.723 |  |  |  |  |
|  | Quadratic | 5.8 | 0.870 |  |  |  |  |
| Female |  |  |  |  |  |  |  |
| F2a | Linear | 90.7 | 0.983 | 3695.75 | 3743.43 | 3757.43 | Okay |
|  | Linear | 9.3 | 0.867 |  |  |  |  |
| F2b | Quadratic | 90.4 | 0.981 | 3755.10 | 3809.59 | 3825.59 | Good |
|  | Quadratic | 9.6 | 0.865 |  |  |  |  |
| F2c* | Linear | 90.7 | 0.981 | 3720.00 | 3771.08 | 3786.08 | Good |
|  | Quadratic | 9.3 | 0.858 |  |  |  |  |
| F3a | Linear | 91.3 | 0.989 | 3801.52 | 3886.66 | 3911.66 | Poor |
|  | Linear | 6.6 | 0.873 |  |  |  |  |
|  | Linear | 2.0 | 0.950 |  |  |  |  |
| White |  |  |  |  |  |  |  |
| W3a | Linear | 55.4 | 0.858 | 5080.68 | 5177.03 | 5204.03 | Poor |
|  | Linear | 37.5 | 0.847 |  |  |  |  |
|  | Linear | 7.1 | 0.866 |  |  |  |  |
| W3b | Quadratic | 52.2 | 0.836 | 5187.47 | 5294.53 | 5324.53 | Good |
|  | Quadratic | 40.0 | 0.855 |  |  |  |  |
|  | Quadratic | 7.8 | 0.847 |  |  |  |  |
| W3c* | Linear | 54.5 | 0.848 | 5096.13 | 5196.05 | 5224.05 | Good |
|  | Linear | 37.9 | 0.849 |  |  |  |  |
|  | Quadratic | 7.6 | 0.859 |  |  |  |  |
| Black |  |  |  |  |  |  |  |
| B3a | Linear | 84.4 | 0.983 | 1046.12 | 1121.90 | 1148.90 | Poor |
|  | Linear | 9.1 | 0.896 |  |  |  |  |
|  | Linear | 6.5 | 0.875 |  |  |  |  |
| B3b | Quadratic | 77.2 | 0.974 | 1075.56 | 1159.76 | 1189.76 | Good |
|  | Quadratic | 14.5 | 0.911 |  |  |  |  |
|  | Quadratic | 8.3 | 0.895 |  |  |  |  |
| B3c | Linear | 84.6 | 0.982 | 1044.15 | 1122.74 | 1150.74 | Poor |
|  | Quadratic | 8.8 | 0.915 |  |  |  |  |
|  | Linear | 6.6 | 0.884 |  |  |  |  |
| B4d | Linear | 76.7 | 0.972 | 1073.93 | 1152.52 | 1180.52 | Okay |
|  | Quadratic | 14.8 | 0.925 |  |  |  |  |
|  | Linear | 8.5 | 0.890 |  |  |  |  |
| B5d* | Linear | 76.9 | 0.973 | 1072.24 | 1153.63 | 1182.63 | Good |
|  | Quadratic | 14.8 | 0.924 |  |  |  |  |
|  | Quadratic | 8.3 | 0.887 |  |  |  |  |

| Sensitivity Analysis |  |  |  |  |  |  |  |
| --- | --- | --- | --- | --- | --- | --- | --- |
| S3a | Linear | 53.7 | 0.821 | 11446.85 | 11554.06 | 11581.06 | Okay |
|  | Linear | 39.9 | 0.833 |  |  |  |  |
|  | Linear | 6.5 | 0.843 |  |  |  |  |
| S3b | Quadratic | 52.9 | 0.820 | 11595.37 | 11714.50 | 11744.50 | Good |
|  | Quadratic | 40.6 | 0.835 |  |  |  |  |
|  | Quadratic | 6.5 | 0.839 |  |  |  |  |
| S3c* | Linear | 53.6 | 0.823 | 11444.59 | 11555.77 | 11583.77 | Good |
|  | Linear | 40.1 | 0.832 |  |  |  |  |
|  | Quadratic | 6.3 | 0.840 |  |  |  |  |

\*Final model selection for each group of models

**Supplementary Figure 1.**
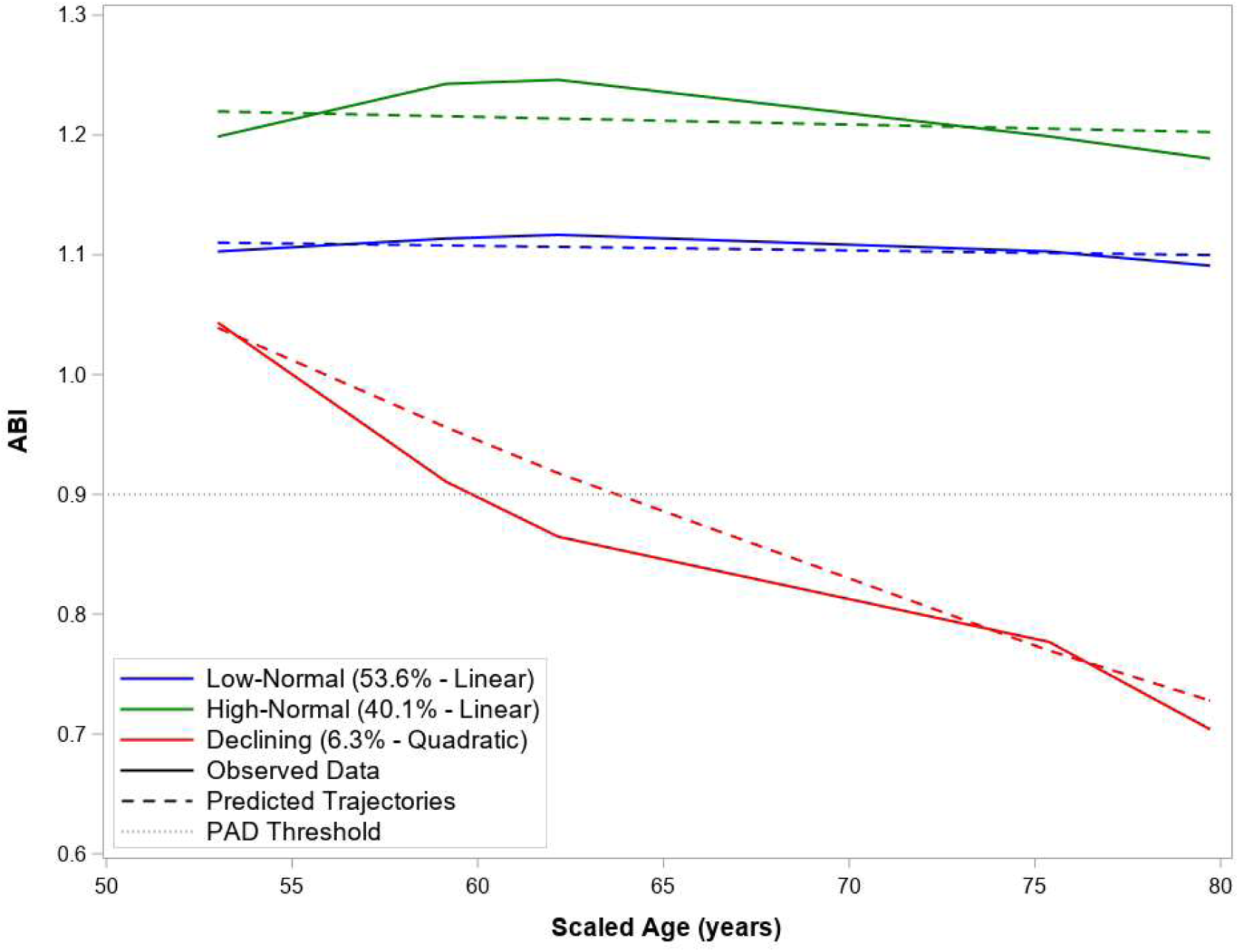
Sensitivity Analysis GBTM Including All Participants with ≥2 ABI Measurements.

